# FAR AWAY FROM HERD IMMUNITY TO SARS-CoV-2: results from a survey in healthy blood donors in South Eastern Italy

**DOI:** 10.1101/2020.06.17.20133678

**Authors:** Josè Ramòn Fiore, Michele Centra, Armando De Carlo, Tommaso Granato, Annamaria Rosa, Michelina Sarno, Lucia De Feo, Mariantonietta Di Stefano, Maria L. D’Errico, Sergio Lo Caputo, Rosella De Nittis, Fabio Arena, Gaetano Corso, Maurizio Margaglione, Teresa Antonia Santantonio

**Author notes:** **Corresponding Author** Jose Ramon Fiore MD, PhD, Department of Clinical and Experimental Medicine, Section of Infectious Diseases, University of Foggia, Ospedali Riuniti University Hospital, Via Luigi Pinto 1 71100 Foggia (Italy).

## Abstract

Here we present results from a survey on anti-SARS-CoV-2 seroprevalence in healthy blood donors from a low incidence COVID-19 area (Apulia region, South Eastern Italy).

Among 904 subjects tested, only in 9 cases (0.99%) antibodies against SARS-CoV-2 were demonstrated. All the 9 seropositive patients were negative for the research of viral RNA by RT-PCR in nasopharyngeal swab.

These data, along with those recently reported from other countries, clearly show that we are very far from herd immunity and that the containment measures are at the moment the only realistic instrument we have to slow the spread of the pandemic.

## INTRODUCTION

Herd immunity is a concept in epidemiology that describes how people can collectively stave off infections if some percentage of the population has immunity to it. When most of a population is immune to an infectious disease, this provides indirect protection, or herd protection, to those who are not immune (1)

There are two ways to achieve herd immunity: a large proportion of the population either gets infected or gets a protective vaccine. Based on early estimates of SARS-CoV-2 infectiousness, it should be likely needed at least 60-70% of the population to be immune to have herd protection (2). However, more recently mathematical models suggest that lower thresholds could be enough to place populations over the herd immunity threshold once as few as 43% (3) or even10-20% of its individuals are immune. (4)

It is therefore important and urgent to evaluate the extent of circulation (and thus of immunity to) Of SARS-CoV-2 in the general populations of affected countries because this information should guide the extent of reduction or increasing of preventive measures such as social distancing etc.

Italy registered the first imported cases of infection in January 31 2020 and after one week the first local case. After that, a dramatic burden of infections was diagnosed: 235.278 cases (with 33.964 deaths) as for **June 8** 2020, with main clusters in Northern Italy. As a whole, 390 infections/100.000 inhabitants. In Foggia (Apulia region, South Eastern Italy) the first case was observed in March the 1st and as for **June 8**, 1162 cases were diagnosed with an incidence of 187 cases/100.000 inhabitants (5)).

Although the total number of diagnosed infections is moderate, we miss clear information regarding the number of individuals in the general population that became immune, possibly acquiring the virus with no or mild symptoms.

Studies on blood donor cohorts are useful to evaluate the prevalence, incidence and natural course of infectious diseases in the general population and may thus help to assess both the viral circulation and the evolution of the COVID-19 outbreak.

We therefore studied a group of healthy blood donors from Foggia province for the presence of IgM and IgG to antibodies to SARS-CoV-2 to examine the circulation of the virus in the general population three months after the local start of the epidemic.

## SUBJECTS, MATERIAL AND METHODS

### SUBJECTS

The main study cohort was composed of blood donors, who were apparently healthy subjects, aged 18-65 years. Exclusion criteria were active infection or medical conditions, recent surgical procedures, stay in endemic areas, reported risk factors for parenterally acquired infections, chronic degenerative conditions, diagnosis of cancer or high risk of cardiovascular events. All donors underwent clinical examination, medical history evaluation and biochemical testing. All subjects should had been free of recent symptoms possibly related to COVID-19, nor had close contact with confirmed cases, symptoms free during the preceding 14 days, nor had contacts with suspected cases.

Each blood donor signed written informed consents allowing for testing for communicable diseases, storing anonymized data and biological materials for diagnostic/research purposes, and use of their anonymous data for clinical research.

A total of 904 blood donors, referring to the Transfusional Center at the “Ospedali Riuniti” University Hospital (Foggia, Italy) were included in the study and subjected to the search for anti-SARS-CoV-2 antibodies, in the period May1-31 2020. In the case of positivity, subjects were re-called and RT-PCR for detection SARS-CoV-2 from nasopharyngeal swabs was performed.

## METHODS

### Detection of anti SARS Cov 2 antibodies

Anti-SARS-CoV-2 IgG and IgM were analyzed by using a chemiluminescent analytical assay (CLIA) commercially available kit (New Industries Biomedical Engineering Co., Ltd [Snibe], Shenzhen, China) performed according to the manufacturers instructions. Reagent wells were coated with recombinant structural protein CoV-S (spike) and e CoV-N (nucleocapside) of SARS-CoV-2 for both IgM and IgG assay. For IgM assay, the microspheres were coated with a monoclonal antibody to capture human IgM followed by the addition of recombinant antigen from virus 2019-nCoV marked with amino-butyl-ethyl-isoluminol (ABEI). The samples, serum or plasma, were diluted by instrument. The relative light units (RLU) detected is proportional to the concentration of IgG/M in sample. An RLU-ratio of the measurement of each sample to the supplied calibrator was calculated. According to manufacturer instructions for the IgG assay arbitrary units of <1 was considered negative, 1.0 to 1.1 borderline and >1.1 positive; for IgM, an AU/mL <0.9 was considered negative, 0.9 to 1.0 borderline and > 1.0 positive.

Manufacturers claimed that the calculated clinical sensitivities of IgM and IgG were 78.65% and 91.21%, respectively, while specificities of IgM and IgG were 97.50% and 97.3%, respectively.

### RT-PCR for detection SARS-CoV-2 from nasopharyngeal swabs

Viral RNA was extracted within 2 hours from sample collection using the STARMag 96 × 4 Universal Cartridge kit with the Microlab NIMBUS IVD instrument according to the manufacturer’s instructions (Seegene Inc. Seoul, Korea). Amplification and detection of target genes (N, E and RdRP) was performed using the commercially available kit AllplexTM 2019-nCoV Assay (Seegene Inc. Seoul, Korea) with the CFX96TM instrument (Bio-Rad, Hercules, CA). Results interpretation was performed with the Seegene Viewer software.

## RESULTS

A total of 904 subjects were enrolled in this study (666 males and 239 females) ranging from 18 to 65 years (Table 1). Among them, 9 tested positive for antibodies to SARS-CoV-2: in 8 cases only IgG antibodies were found while in 1 case both IgG and IgM were detected, with a percentage of positivity in the blood donor population of 0.99 %.

**TABLE 1:** Detection of antibodies to SARS Cov 2 in 904 healthy blood donors according to sex and age

|  | **n°** | **Positive n°** | **%** |
| --- | --- | --- | --- |
| **Total** | 904 | 9 | 0.99 |
| **Males** | 665 | 5 | 0.75 |
| **Females** | 239 | 4 | 1.6 |

| **Ages (range)** | **n°** | **n°** | **%** |
| --- | --- | --- | --- |
| **Group I (18 – 25)** | 112 | 1 | 0.89 |
| **Group II (26 – 35)** | 195 | 4 | 2.0 |
| **Group III (36 – 45)** | 202 | 1 | 0.49 |
| **Group IV (46 – 55)** | 246 | 0 | 0 |
| **Group IV (56– 65)** | 149 | 3 | 2.0 |

No statistical differences were observed in the rate of antibody detection according to the sex and the age.

None of these subjects had clinical signs or symptoms and laboratory parameters alterations; all of them tested negative for RT-PCR research of SARS-CoV-2 RNA in two nasopharyngeal swabs performed on two consecutive days.

## DISCUSSION

Herd immunity is the resistance to the spread of a contagious disease that results if a sufficient proportion of a population is immune due to natural infection or mass vaccination.

Because an effective COVID-19 vaccine is not yet available, herd immunity can be established if a large proportion of immune persons exist in a population to confer indirect protection from infection to susceptible individuals. The breadth of the barrier required to achieve herd immunity depends in large part on viral spread and infectivity. In the nomenclature of epidemiology, the basic <u>reproduction number</u> or R0 and the classical formula for calculating a herd immunity threshold is 1—1/R0. The higher the R0, the higher the threshold required for achieving herd immunity. Other important factors in calculating herd immunity thresholds include the number of social interactions and their durations, innate differences in individual immune responses, and <u>divergent exposures</u> to the infectious microbe (1)

People who recover from a COVID-19 coronavirus infection, at least for some time, develop immunity to the virus (6).

The disease-induced herd immunity threshold for SARS-CoV-2, according to various epidemiologists, is believed to be around 60 to 70 percent (2) More recently, some researchers have reported lower thresholds: from 43% to just 10 to 20% of the population (3, 4), suggesting that we could reach herd immunity thresholds by natural infections in the setting of COVID-19 pandemic.

Recently, data from Spain, France and Italy (7–9) countries that adopted strict lockdown measures indicate a very low seroprevalence in the general population (4.4%, 5% and 7.1%, respectively). Noteworthy, even in Sweden, a country that decided for a herd immunity strategy, with very light restrictions on daily life, antibodies to SARS-CoV-2 were detectable in only 7.5% of the general population in Stockholm (10).

In other geographical regions, serosurveys in healthy individuals demonstrated a very low rate of positivity for SARS-CoV-2 (11–15).

In our study, we confirm a low rate of antibodies against SARS-CoV-2 on a group of blood donors from a geographical region with a moderate incidence (187 cases/100.000 inhabitants vs the national data of 390 infections/100.000 inhabitants).

A limitation of our study is that the enrolled population (18-65 years old) is representative of only a part of the general population, since in Italy 22% of the individuals is aged >65 years (16) However, presented data are relevant in that they refer to the more acting/interacting group of the population, also important from a productive point of view.

Certainly, we are far away from herd immunity and even from the more optimistic projections of threshold (10 to 20% of the population) to adopt more relaxed strategies.

In conclusion, strengthening herd immunity to control the COVID-19 epidemic is not a viable option as large numbers of people are expected to become infected and many may die from COVID-19. Preventive measures, including physical distancing, remain essential to contain the spread of infection until herd immunity can be safely acquired with the vaccine. Health authorities should take in account these considerations when facing the public health measures to be adopted throughout the COVID-19 transition phases.

## Data Availability

at request

